# COVID-19 modulates pregnancy outcomes

**DOI:** 10.1101/2025.06.09.25328575

**Authors:** Maria Laura Gabriel-Kuniyoshi, Sérgio Nery Simões, Lucas Pinheiro Badaró Moreira, Juliana Yuri Kanezaki de Souza, Thaina Oliveira Nogueira, Tatiane Maria Angelo Catharini, Jackeline A. Della Torre, Bianca S. Leoni, Kethellen E. Biancolin, Silvia Maria Ibidi, Rossana Pulcineli Vieira Francisco, Ester Cerdeira Sabino, Caroline Camilo, Gisele Rodrigues Gouveia, Alexandra Brentani, David Correa Martins, Helena Brentani

## Abstract

**Introduction:** The COVID-19 pandemic exposed many pregnant individuals to SARS-CoV-2. Literature suggests a link between gestational COVID-19 and adverse gestational outcomes. However, among all the factors that could impact pregnancy, COVID-19 is not the sole determinant, mainly in vulnerable populations. We hypothesized that COVID-19 could act as a moderator of some environmental exposures and pregnancy outcomes.

**Methods:** To investigate this, our sample consisted of 120 pregnant individuals with or without a history of SARS-CoV-2 infection (moderator) and delivery complications (outcome). eXplainable Artificial Intelligence (XAI) was used to select the exposures from fifteen variables: trait and state anxiety, perceived stress in the past month, lifetime and gestational stressful events, body mass index, number of prenatal appointments, the violent crime rate in their area, belonging to a marginalized ethnic group, paternal and maternal age, income, education level, and employment. A moderation analysis evaluated whether COVID-19 exerted a moderator effect between the exposures and outcome.

**Results:** XAI showed four variables that were associated with both pregnancy complications and COVID-19: trait anxiety, state anxiety, paternal age, and maternal age. These four variables were set as exposures in moderation analyses where COVID-19 was a moderator and gestation complications were the outcomes. COVID-19 interaction with the exposures was significant.

**Discussion:** Our research suggests that being infected by SARS-CoV-2 exacerbates the effects of anxiety and parental age on susceptibility to adverse pregnancy outcomes. In other words, COVID-19 is a moderator for these variables, which should be accounted for in future studies.

**Significance:** *What’s Known on This Subject?:* Infection by the virus SARS-CoV-2 causes the COVID-19 disease, which led to a worldwide pandemic. Pregnant people with COVID-19 have an increased risk of delivery complications such as preeclampsia. Other variables, such as mental health, also influence pregnancy outcomes.

*What This Study Adds?:* Our sample showed no significant relationship between COVID-19 and delivery complications. However, COVID-19 interacted with anxiety and parental age to potentiate their effect on delivery complications. We suggest that COVID-19 worsens the challenges for young and anxious parents.

## Introduction

The COVID-19 Pandemic, caused by the SARS-CoV-2 virus, is one of the most significant events in recent history. It has led to over 775 million confirmed cases and 7 million deaths worldwide (World Health Organization, 2024). These fatalities keep occurring even after the World Health Organization declared the end of the public health emergency. Acute illness and death are not the only consequences of COVID-19: it may also cause long COVID-19 (Davis et al., 2023), increased risk of neuropsychiatric conditions (Taquet et al., 2022), and adverse impacts on the children of infected pregnant people (Ciapponi et al., 2021). As for the latter, studies link COVID-19 during pregnancy to adverse outcomes such as low neonate weight, preterm birth, and pregnancy complications (Allotey et al., 2020; Ciapponi et al., 2021).

However, COVID-19 is not the sole determinant of pregnancy outcome. Among the many variables relevant to pregnancy outcomes, we can cite maternal age: mothers older than 40 years have 132% higher odds of experiencing a miscarriage (Khalil et al., 2013). Another example is mental health — mothers with anxiety and depression face 17% higher odds of adverse pregnancy outcomes (Rejnö et al., 2019). To further complicate, some factors that influence pregnancy outcomes also affect COVID-19 susceptibility, including the above-mentioned age and mental health (Monod et al., 2021; Yang et al., 2020). This interplay creates a complex landscape of COVID-19, additional exposure variables, and pregnancy outcomes.

These relationships can take several forms. Confounders are variables influencing both exposure and outcome (Morrow et al., 2022). Mediators, in contrast, intermediate the causal pathways between an exposure and an outcome (Morrow et al., 2022). Finally, moderators affect the strength or even the direction of the relationship between an exposure and an outcome; for instance, the relationship between mental health and gestational outcomes might differ between people who had COVID-19 or not.

Here, we hypothesize that COVID-19 could act as a moderator between exposures and outcomes, for instance, the relationship between mother’s mental health and gestational outcomes might differ between people who had COVID-19 or not. With eXplainable Artificial Intelligence (XAI), we evaluated fifteen socioeconomic, psychological, and health variables, selecting those associated with COVID-19 infection. Next, also using XAI, we verified how the selected variables and COVID-19 contribute to pregnancy outcomes, further refining the variable selection. A moderation analysis investigated whether COVID-19 is a moderator in the association between the selected variables and the gestational outcome. As we could not find an association between COVID-19 and pregnancy outcomes, we discarded the hypothesis of a mediator role of COVID-19. Our results indicate that gestational COVID- 19 intensifies the effects of maternal state anxiety, maternal trait anxiety, maternal age and paternal age on the risk of adverse pregnancy outcomes.

## Methods

### Overview, Strategy and Ethical Considerations

This research focuses on whether COVID-19 moderates or mediates associations between environmental exposures and gestational outcomes. Our first task is to identify the relevant exposures associated with COVID-19 and delivery complications, using XAI. We also used XAI to evaluate the association between COVID-19 and delivery complications. We could not confirm the association between COVID-19 and delivery outcomes, thus we did not test a mediation relationship.

#### Ethics Statement

The Research Ethics Committee of the Hospital Universitário da Universidade de São Paulo (HU-USP) approved this study on 02/26/2021 (protocol 40160520.7.0000.0076). Our research conforms with the Brazilian General Personal Data Protection Law (13709/2018), which guarantees data privacy rights, and the World Medical Association Declaration of Helsinki. All the participants signed a consent form. No identifying information is published in any source.

#### Study design and participant recruitment

This cross-sectional study recruited 353 pregnant females at delivery, in the HU-USP, between March 2021 and March 2022. The HU-USP is located in São Paulo City, in the Butantã district, which is characterized by socioeconomic heterogeneity with slums, a university town, and upper-class neighborhoods. If the patient signed the consent form, we collected their blood and administered a questionnaire to assess social, physical, and mental health variables. Medical records were evaluated to determine mothers and neonates’ conditions. The inclusion criteria were pregnant people who reside in the São Paulo Metropolitan Area, gave birth at HU-USP, and have agreed to participate in the study. Exclusion criteria were: unaccompanied minor parents, deliveries in which the 5th minute Apgar score was under 7, and cases of congenital malformation.

### Exposure, outcome and COVID-19 definitions

#### Outcome

The outcome used was the presence of delivery complications registry by the Hospital Staff in the medical records (0 = no complications, 1 = reported complications). These include acute fetal distress, eclampsia, preeclampsia, cephalopelvic disproportion and instrumental delivery. We opted for this broader outcome since the frequency of specific factors, such as preeclampsia, was too small to undergo analysis in our sample. Cesareans are not included as an adverse outcome due to the cultural preference for this delivery type in Brazil (Knobel et al., 2020).

#### COVID-19

Our moderator was whether the patient had COVID-19 (group COVID, value=1) or not (group CONTROL, value=0). To determine this value, immunoassays double-checked the presence of anti-SARS-CoV-2 antibodies in the patient’s blood. We collected maternal peripheral blood at delivery and stored it at room temperature for up to 6 hours. The blood was centrifuged at 3000g for 10 minutes and stored at -20°C. The first immunoassay detected general anti-SARS-CoV-2 antibodies using the SARS-CoV-2 Ag Rapid Test (Guangzhou Wondfo Biotech, Huangpu District Guangzhou, China). The second was specific to IgG antibodies for nucleocapsid proteins (Elecsys®, Roche Diagnostics International Ltd, Rotkreuz, Switzerland) to be able to distinguish SARS-CoV-2 infection from immunization by most vaccines. The only vaccine that cannot be distinguished from a proper infection is CoronaVac (Sinovac Biotech Co. Ltd, Haidian District, Beijing, China), so we used the patients’ vaccination history to account for this factor.

#### Exposure variable descriptions

Fifteen variables covered demographics, gestational health, and maternal mental health **(Online Resource Table 1)**. We selected only 15 variables to avoid the curse of dimensionality, keeping a proportion of six samples per variable. The variables correlated less than 70% among themselves **(Online Resource Figure 1)** and had less than 15% missing data.

**Fig 1.**
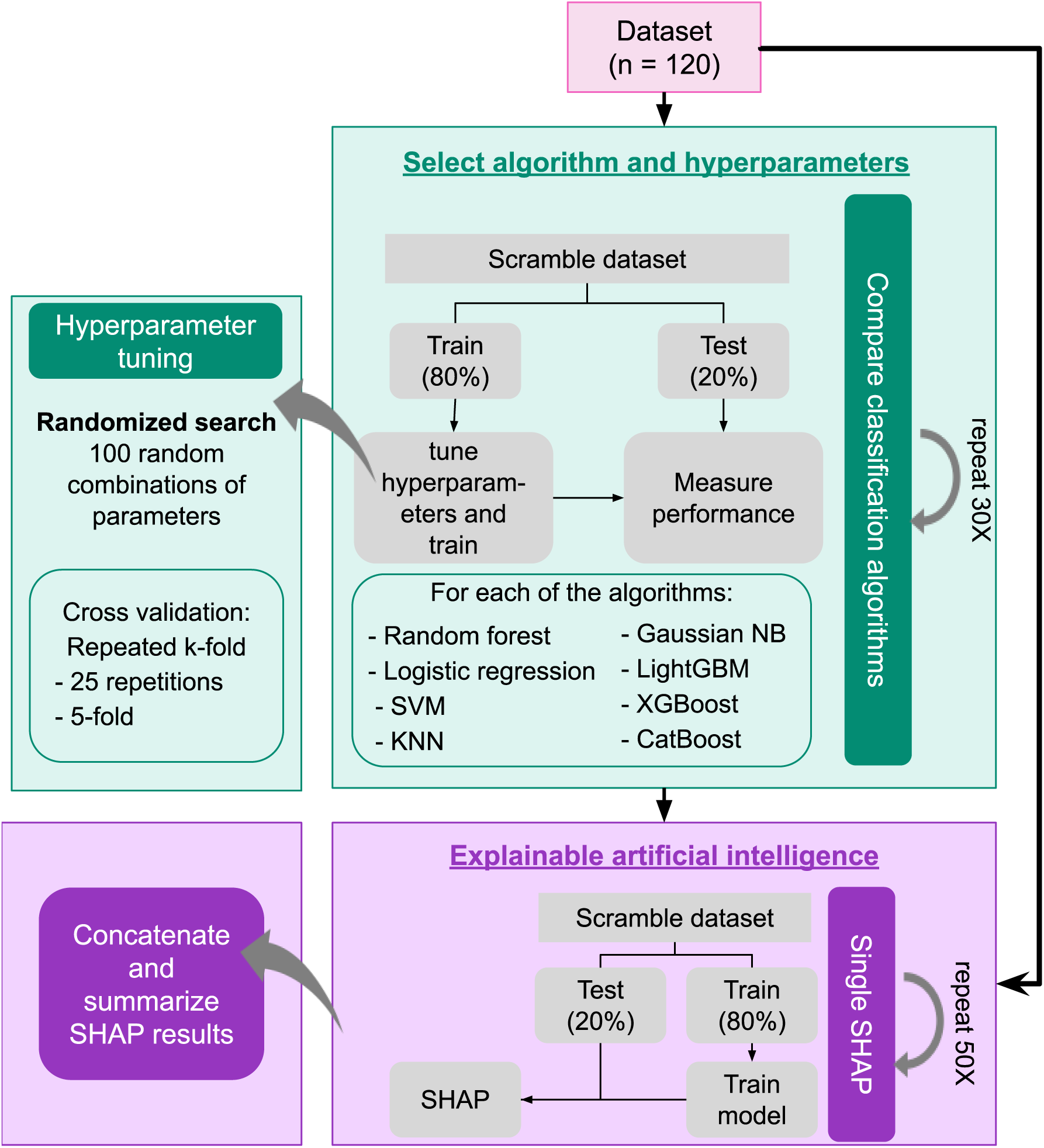
eXplainable Artificial Intelligence (XAI) workflow for predicting whether the patient had COVID-19. The process was divided into selecting the best algorithm and hyperparameters, and model explanation. Loops of repeated cross-validation aimed at compensating the small sample size. While the top part (green) was applied to COVID-19 only, the bottom part (purple) was applied to both COVID-19 and delivery complications. Adapted from Costa et al (Costa et al., 2022)

Interviews at the hospital during the postpartum stay were used to collect demographic and health data. Properly translated instruments, which have already been validated in a Brazilian population, were also administered during the interview. For mental health, the State-Trait Anxiety Inventory (STAI) (Biaggio et al., 1977) measured both trait and state anxiety. Trait anxiety is a long-lasting state that increases the predisposition to anxious behavior and feelings, while state anxiety is a transient status in response to current events. The Perceived Stress Scale (ten-item version - PSS10) (Luft et al., 2007) measured stress one month before delivery, while two in-house questionnaires assessed whether stressful events occurred during lifetime and pregnancy. For the environment, we retrieved the violent crime rate in their district of residence (São Paulo Public Security Secretariat, 2024). **Online Resource Table 1** provides details on how we obtained each variable.

### Data processing and machine learning

Patients with over 20% missing data were removed. For the remaining dataset, 2.77% of the values were still missing, so Scikit-learn 1.2.1 KNNImputer() (Pedregosa et al., 2011) replaced them with the mean value from the three nearest neighbors. Such a number of neighbors was selected by machine learning and data distribution experiments **(Online Resource Figure 2)**. Z-score standardized variables were used for subsequent analysis. The processing occurred on Pandas 1.5.3.

**Fig 2.**
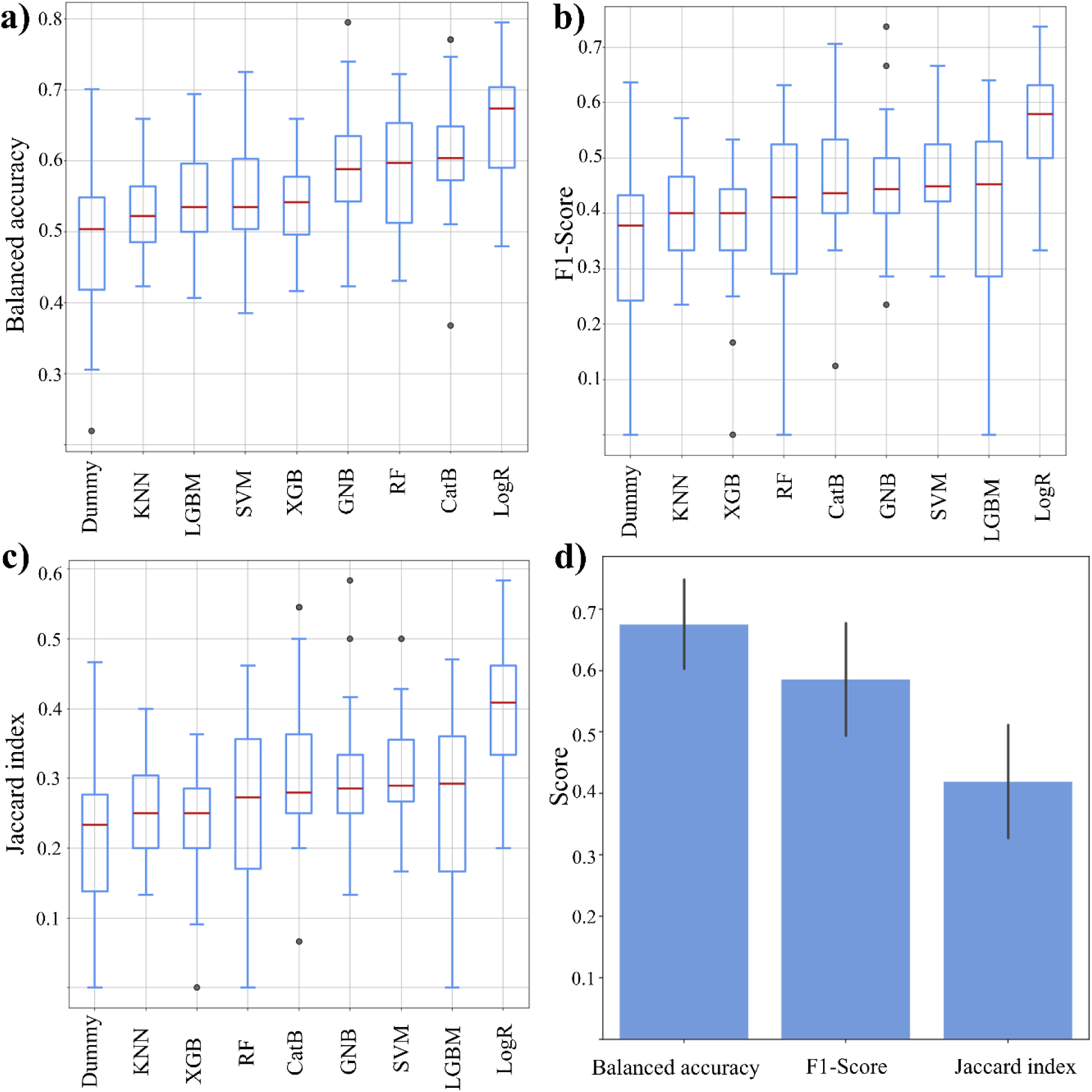
Comparison of models predicting whether the pregnant person had COVID-19. **a-c)** Accuracy, F1-score, and Jaccard index for the models: support vector machine (SVM), k-nearest neighbours (KNN), Gaussian Naïve-Bayes (GNB), LightGBM (LGBM), XGBoost (XGB), CatBoost (CatB), logistic regression (LogR) and random forest (RF), as well as a random baseline (Dummy). Boxes represent the Q2 and Q3 quartiles, the line represents the median, and the whiskers represent 150% of the interquartile range. **d)** Performance metrics for the selected hyperparameter combination, from the LogR algorithm

#### XAI for predicting COVID-19

To identify which exposures could be moderated by COVID-19, we performed an XAI approach to find variables associated with the COVID-19 group. The first step was to find the best algorithm and hyperparameters for the machine learning analysis. Our methodology had multiple cross-validation steps to avoid overfitting **(Figure 1)** (Costa et al., 2022). We executed thirty comparisons of eight classification algorithms: random forest (Scikit-learn v1.3.0), logistic regression (Scikit-learn v1.3.0), LightGBM (v4.3.0) (Ke et al., 2017), XGBoost (v2.0.3) (Chen & Guestrin, 2016), CatBoost (v1.2.3) (Prokhorenkova et al., 2019), support vector machine (SVC function from Scikit-learn v1.3.0), k-nearest neighbors (Scikit-learn v1.3.0), and Gaussian Naïve-Bayes classifier (Scikit-learn v1.3.0). Each comparison had a different shuffle of samples, with a stratified test-train split to account for the unbalanced classes. A randomized search tested 100 possibilities of hyperparameter combinations, using the Scikit-learn function RandomizedSearchCV() and the hyperparameter space shown in **Online Resource Table 2**.

After choosing the algorithm and hyperparameters combination, we performed 250 SHapley Additive exPlanations (SHAP) (Lundberg & Lee, 2017) v0.42.1 to identify the most important variables. Each experiment had different scrambles of samples generated by 25 repeats of 5-fold stratified cross-validation. The several SHAP loops compensate for the small sample size. The 250 experiments were collapsed to provide an average view.

#### Clustering Analyses

We clustered the exposure variables by their SHAP values to select the most unique contributions to the model predictions. We considered the median SHAP values of the 250 runs. We performed the hierarchical clustering analysis with the function clustermap() of the Seaborn library v0.13.2 in Python, considering Euclidean distance and Ward’s method. The cluster cut-off was seven.

#### XAI for predicting exposure-outcome associations

A second XAI analysis examined the relationship between potential exposures and delivery complications. This analysis focused on the seven most relevant variables in the COVID-19 SHAP values clustering. We also included COVID-19. To ensure comparability, we applied the same algorithm and hyperparameters selected in the COVID-19 XAI. Thus, we followed only the methods illustrated in the bottom portion of **Figure 1**.

### Moderation Analyses

Moderation analyses were performed in R 4.2.1 programming language with the lavaan v0.6.18 package (Rosseel, 2012), using a diagonally weighted least squares (DWLS) estimator and bootstrap with 1000 draws.

### Hardware and software specifications

The hyperparameter tuning was performed in the Santos Dumont supercomputer, using 21 nodes with 64GB of RAM each, Intel Xeon CPU E5-2695 v2 processor, operating on Linux system, on Python 3.12.2. The tuning took 137h48m of CPU time. The data processing, final model training, SHAP analysis, and moderation Analysis were performed on a Dell laptop with 16GB of RAM, with an 11th Gen IntelCore i7 processor, operating on Windows 11 system, Python 3.9.16. The repository https://github.com/marialgk/PregnantCOVID/ contains our scripts.

## Results

### Sample description

Between 2021 and 2022, the HU-USP hospital had an average of 1904 births per year, and 353 patients consented to participate in the study. When evaluating COVID-19, we discarded 152 who missed COVID-19 data and removed 80 patients with inconclusive COVID-19 evaluations. After removing patients with over 20% missing total data, the sample size was 120 **(Online Resource Figure 3)**. Seven patients missed records on delivery complications, resulting in 113 samples, of which 42 were COVID and 71 were CONTROL. **Online Resource Table 3** shows the descriptive statistics of the final sample. **Online Resource Table 1** indicates the percentage of missing data.

**Fig 3.**
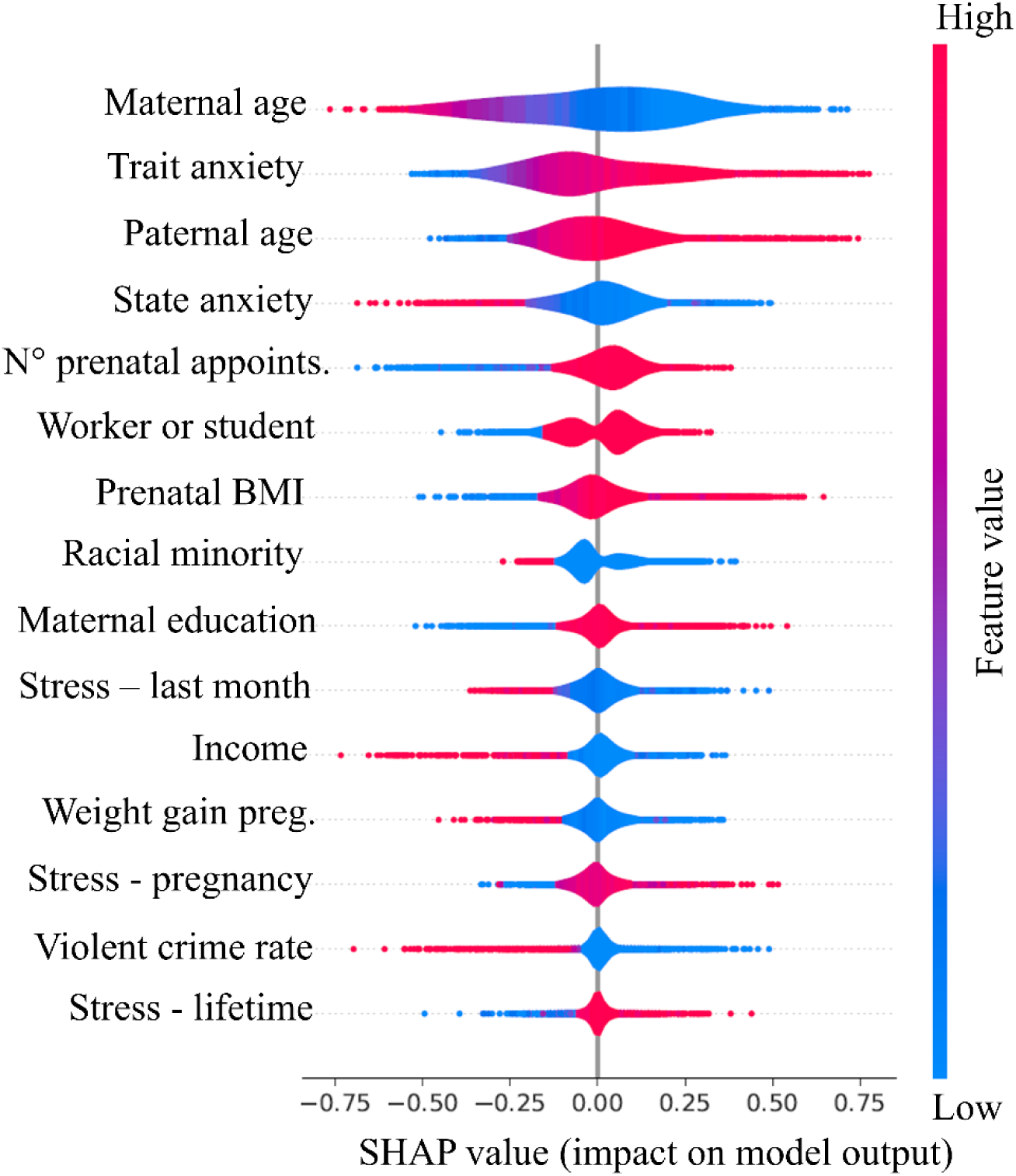
eXplainable Artificial Intelligence (XAI) for a logistic regression model that predicts whether the pregnant person has previously had COVID-19. This figure derives from the concatenation of 250 SHAP experiments. The y-axis represents the predictor variables ordered by explanatory importance. The color scale represents the variable values standardized by z-score, where red is closer to 1 and blue is closer to -1. The x-axis represents the SHAP value, which indicates how much the variable contributes to the class prediction (positive values for the COVID-19 class, negative values for the CONTROL class). N° prenatal appoints. = number of prenatal appointments; Weight gain preg. = weight gain during pregnancy

### Age and anxiety factors predict COVID-19 in pregnant people

An exploratory analysis involving a pairwise distribution of nine predictor variables **(Online Resource Figure 4)** did not identify any visible pattern distinguishing the groups. Therefore, we tested several machine learning algorithms to uncover the most representative features of our data. We tuned the hyperparameters and compared the performance metrics of eight classifier algorithms **(Figure 2)**. Logistic regression classifiers had the highest median balanced accuracy (median 67.3%), F1-score (median 0.580), and recall (median 66.7%). The best hyperparameters were liblinear solver, balanced class weight, L2 penalty, and learning rate of 1. Although our tests suggested a linear model, SVM with linear kernel underperformed, which was confirmed by additional tests **(Online Resource Figure 5)**.

**Fig 4.**
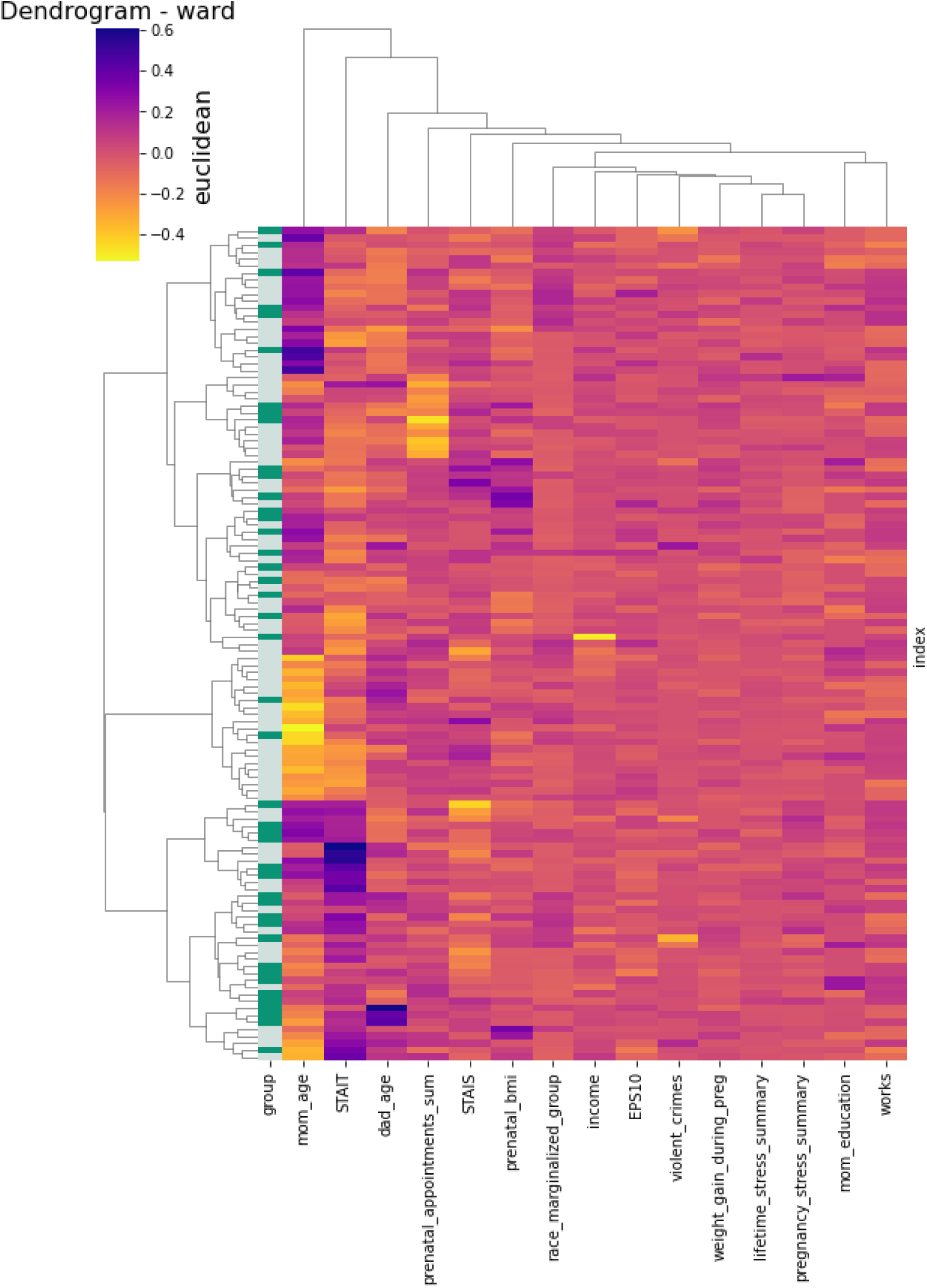
Hierarchical clustering of the median SHAP values. The clustering considered Euclidean distance and Ward’s method. Rows represent patients while columns represent the predictor variables. The red-yellow-blue gradient represents the median SHAP value. The green color code represents the real target value, i.e. whether the patient is from the COVID or CONTROL group.

**Fig 5.**
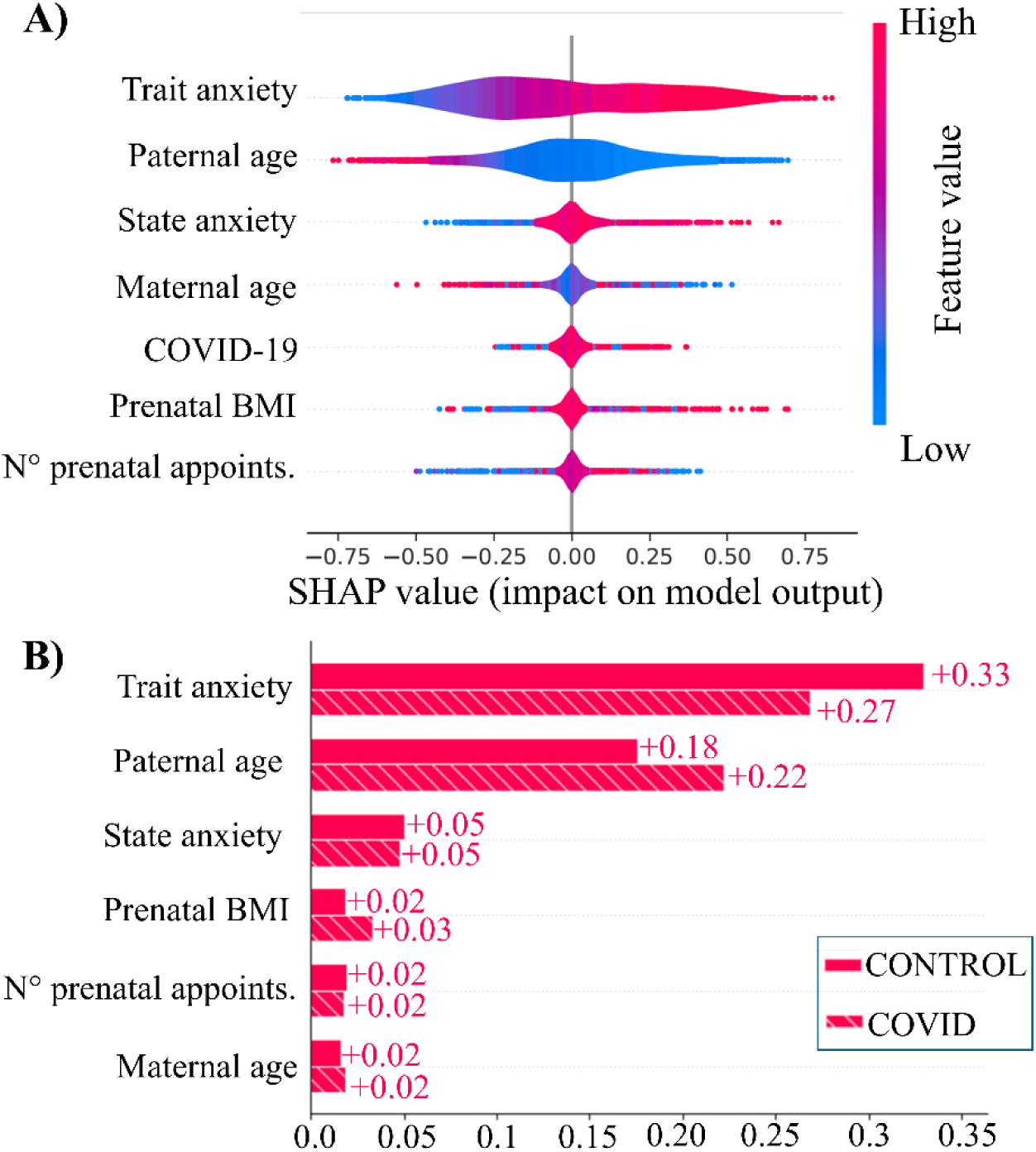
eXplainable Artificial Intelligence (XAI) for a logistic regression model that predicts whether the medical staff recorded any delivery complications at delivery. **A)** Concatenation of 250 SHAP experiments. The y-axis represents the predictor variables ordered by explanatory importance. The color scale represents the variable values standardized by z-score, where red is closer to 1 and blue is closer to -1. The x-axis represents the SHAP value, which indicates how much the variable contributes to the class prediction (negative values = no recorded delivery complications; positive values = reported delivery complications). **B)** Result of a single SHAP experiment, showing the SHAP values separately for patients of the COVID and the CONTROL classes. The x-axis represents the mean global importance across the samples

The chosen models were analysed with XAI, which showed that the most impactful variables were maternal age, trait anxiety, paternal age, state anxiety, and the number of prenatal appointments **(Figure 3)**. A classification towards the COVID group seemed to relate to a younger maternal age, older paternal age, higher trait anxiety, lower state anxiety, and having attended more prenatal appointments.

Then, we used SHAP values, which indicate how much the variables influenced the prediction, to run a hierarchical clustering. This way, we would understand which variables are most uniquely related to COVID-19 (**Figure 4)**. Considering a threshold of seven clusters, six clusters contained a single variable, and one cluster was formed by the remaining nine variables. The six variables that formed a cluster of their own were: maternal age, trait anxiety, paternal age, number of prenatal appointments, state anxiety, and prenatal body mass index (BMI). The red dotted lines represent the Euclidean value thresholds that we consider as most relevant for defining groups for our analysis

### Age, anxiety and COVID-19 in a model to predict delivery complications

We selected the six variables that formed their own cluster in the hierarchical clustering, plus COVID-19. They were considered predictors for the subsequent classification experiments, where the target was whether there were any delivery complications. We used the same algorithm and hyperparameter space as the COVID-19 analysis **(Figure 5A)**.

We now contrast the results of the XAI experiments with COVID-19 or delivery complications **(Figures 3 and 5)**. In both analyses, the variables that mostly influenced the prediction were trait anxiety, state anxiety, paternal age, and maternal age. Paternal age and state anxiety had an inverse direction in the two experiments: while older paternal age and lower state anxiety predicted COVID-19, younger paternal age and higher state anxiety predicted delivery complications. In contrast, trait anxiety had the same direction as both COVID-19 and delivery complications: high trait anxiety predicted COVID-19 and delivery complications.

Now we analyze the meaning of COVID-19 as a predictor of delivery complications. COVID-19 had a low impact on the classifier prediction **(Figure 5)**. This raised the hypothesis that, in our sample, COVID-19 was insignificant as a predictor itself, but it still had a hole as a moderator of other exposures. To further understand this effect, we performed a single SHAP experiment and observed whether the exposures had varying importance for the COVID and the CONTROL groups **(Figure 5B)**. Paternal age and trait anxiety predict delivery complications differently in the CONTROL and COVID groups, supporting the hypothesis of a moderator effect.

### COVID-19 exacerbates the effects of maternal anxiety and parental age on delivery complications

The previous experiments selected Trait anxiety, State anxiety, maternal age and paternal age as exposure variables to the delivery complications outcome. Moderation analysis tested this outcome-exposure relationship under the moderation of COVID-19 **(Figure 6, Online Resource Table 4)**. In any of the four models, there was no significant individual association between COVID-19 and delivery complications, which indicated that direct or mediation effects of COVID-19 could not be observed in our data, so we discarded these hypotheses. However, there was a significant moderation effect: previous COVID-19 exacerbated the association between exposures and outcomes **(Online Resource Figure 6)**. This suggests that COVID-19 moderates trait and state anxiety, maternal age and paternal age interactions with delivery complications.

**Fig 6.**
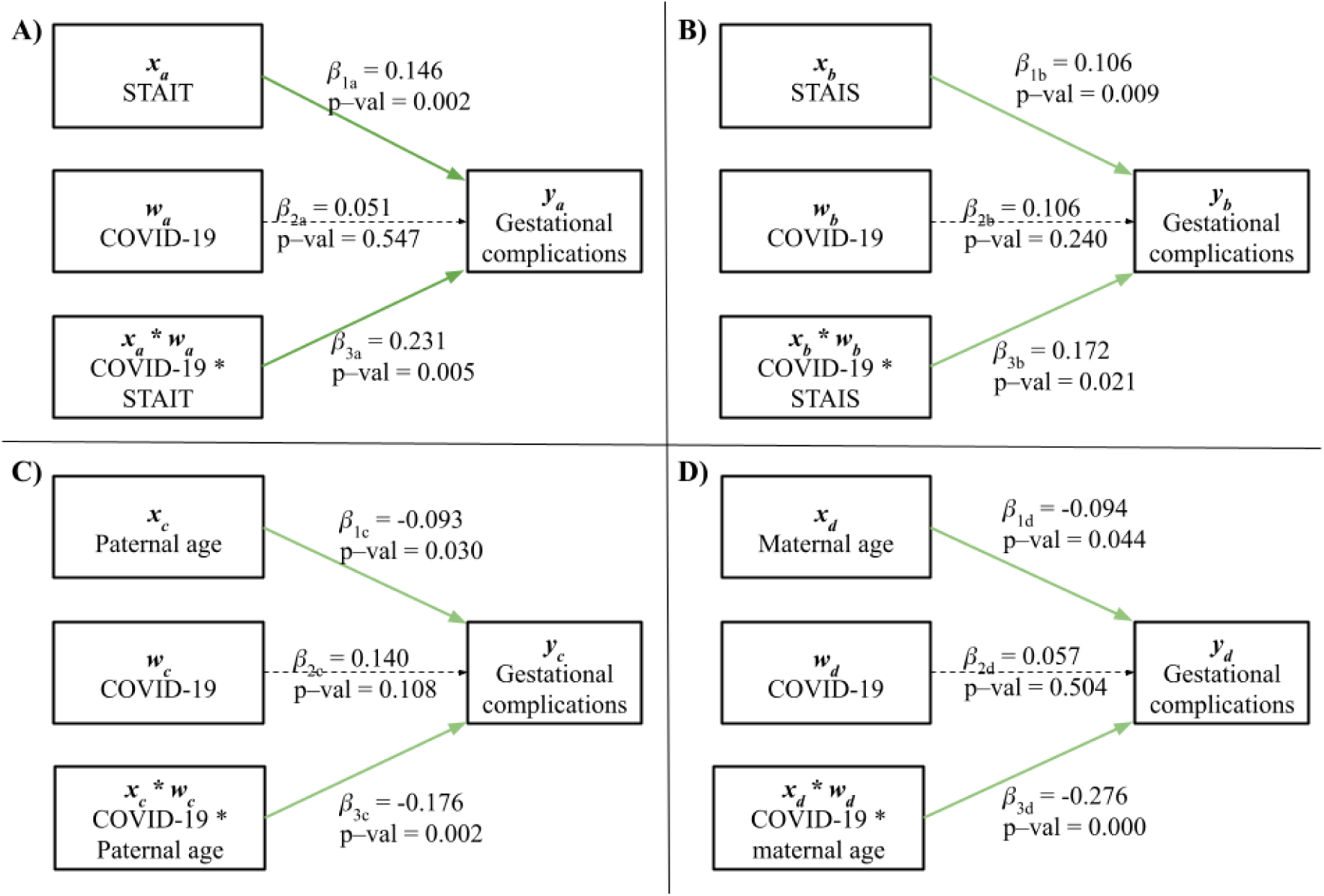
Parameter estimates and p-values in the four moderation models. While the exposure varied across the four analyses, the moderator always was COVID-19, and the outcome was delivery complications. Green arrows indicate statistical significance, while dotted black arrows indicate no significance. STAIT = trait anxiety; STAIS = state anxiety

To get further insight into the combined effects of the four exposures, we tested a multivariate model that included all of them simultaneously. However, due to the high number of parameters (55) relative to the limited sample size (113), the model was saturated (just identified), leaving no degrees of freedom for assessing model fit.

## Discussion

This study has used machine learning and moderation analysis to investigate the relationship between COVID-19, pregnancy outcomes, and environmental exposures. Unlike previous reports (Allotey et al., 2020; Ciapponi et al., 2021), we could not identify a direct effect of COVID-19 infection on the risk of adverse pregnancy outcomes. However, we found a significant moderation effect of COVID-19 on state anxiety, trait anxiety, paternal age, and maternal age. This section will discuss the results and purpose hypotheses on the mechanisms underlying the identified associations.

We found an association between anxiety and the occurrence of pregnancy complications, which was already known to the scientific community: studies conducted in several populations have shown that various types of anxiety relate to outcomes such as preeclampsia, low birth weight, vaginal bleeding and preterm labor (Hosseini et al., 2009; Rejnö et al., 2019). The complete mechanism remains to be clarified, but it is likely related to changes in stress hormone levels, such as a cortisol increase and inflammation markers (Johnson & Slade, 2003). As a novel result, our study shows this association being exacerbated in patients who had COVID-19. Supporting our hypothesis, previous studies have shown that SARS-CoV-2 infection increases the odds of having moderate to severe anxiety by more than two-fold (Alacevich et al., 2023). Interestingly, one study has linked long COVID to lower cortisol levels (Klein et al., 2023), but this result remains controversial (Alaedini et al., 2024).

Another result is the relationship between parental age, delivery complications, and COVID-19. In our study, the younger the parents, the higher the odds of having adverse maternal outcomes. This may be due to our data set including parents as young as 15. Indeed, the two groups with an increased risk of adverse pregnancy outcomes are adolescents and people over 35 (Cavazos-Rehg et al., 2015; Khalil et al., 2013). Regarding the moderator effect, COVID-19 exacerbated this association between age and outcomes. Therefore, our results suggest that SARS-CoV-2 infection piles up the challenges faced by young parents. Especially in developing countries, the COVID-19 pandemic has been linked to a rise in adolescent pregnancy and poorer reproductive health, which could contribute to risky and unplanned pregnancies (Okeke et al., 2022; Zulaika et al., 2022).

Nevertheless, the findings have some limitations. Our sample was small and unbalanced, consisting of 120 samples and an overrepresentation of the CONTROL group. For logistic regression, which was used in our study, the recommended sample size ranges from 10 to 50 samples per independent variable (Bujang et al., 2018; Peduzzi et al., 1996). Our experiment had only six samples per variable, but to address this issue we used a series of cross-validation loops. As a second limitation, while our COVID-19 test shows whether there was an infection during pregnancy, it cannot indicate when it happened. This is critical because the infection timing can affect fetal development differently (Ayed et al., 2022). Despite those limitations, our study proposes new ideas and opens opportunities for future research.

A potential question is whether SHAP analysis is necessary when using logistic regression, as the regression coefficients already provide insights into predictor influence. For linear models like logistic regression, SHAP values can be directly approximated from the model coefficients (Lundberg & Lee, 2017). Despite this, combining SHAP and logistic regression enhances interpretability and visualization. SHAP accounts for collinearity and interactions, which regression coefficients do not represent explicitly. Also, SHAP’s local interpretability feature makes it more intuitive to inspect an individual prediction than with a logistic regression model. Although not our case, SHAP is useful when variables have different units and are unstandardized: logistic regression coefficients depend on variable magnitudes, while SHAP provides scale-free values. SHAP analysis has been widely applied to logistic regression models in the literature (Ahmed et al., 2024; Huang et al., 2025), including the documentation of the SHAP package.

In conclusion, we have found that SARS-CoV-2 infection, at an undetermined point of gestation, increases the effect of maternal anxiety and parental age on pregnancy outcomes. These relationships, which have not been acknowledged in previous studies on the effects of COVID-19 in pregnant people, should be considered in subsequent works. Future research may confirm our results with a larger sample and investigate the underlying mechanisms.

## CRediT authorship contribution statement

**Maria Laura Gabriel-Kuniyoshi:** Data Curation, Formal Analysis, Methodology, Software, Visualization, Writing – Original Draft, Writing – Review & Editing;

**Sérgio Nery Simões:** Supervision, Methodology, Software, Visualization, Writing – Original Draft, Writing – Review & Editing;

**Lucas P. B. Moreira:** Software, Formal Analysis;

**Juliana Yuri Kanezaki de Souza:** Software, Formal Analysis;

**Thaina Oliveira Nogueira:** Data Curation, Validation, Investigation;

**Tatiane Maria Angelo Catharini:** Data Curation, Investigation;

**Jackeline A. Della Torre:** Data Curation, Investigation;

**Bianca S. Leoni:** Data Curation, Investigation;

**Kethellen E. Biancolin:** Data Curation, Investigation;

**Silvia Maria Ibidi:** Data Curation, Investigation;

**Rossana Pulcineli Vieira Francisco:** Investigation, Resources;

**Ester Cerdeira Sabino:** Investigation, Resources;

**Caroline Camilo:** Formal Analysis, Investigation, Project Administration, Visualization, Writing – Original Draft, Writing – Review & Editing;

**Gisele Rodrigues Gouveia:** Investigation, Data Curation, Methodology, Project Administration, Writing – Original Draft, Writing – Review & Editing;

**Alexandra Brentani:** Conceptualization, Project Administration, Supervision, Resources, Funding Acquisition, Writing – Review & Editing;

**David Correa Martins-Jr:** Conceptualization, Methodology, Supervision, Visualization, Writing – Original Draft, Writing – Review & Editing;

**Helena Brentani:** Conceptualization, Project Administration, Supervision, Resources, Funding Acquisition, Methodology, Writing – Original Draft, Writing – Review & Editing.

All authors approved the manuscript.

## Data availability

All the results are available in this manuscript and its supplementary material. The anonymized and processed dataset, as well as the scripts used for the analysis, are available at https://github.com/marialgk/PregnantCOVID/. More information can be obtained with the corresponding author upon request.

## Abbreviations

BMI: Body Mass Index
COVID-19: Coronavirus Disease 2019
DWLS: Diagonally Weighted Least Squares
HU-USP: Hospital Universitario of São Paulo University
PSS10: Perceived Stress Scale (ten-item version)
SARS-CoV-2: Severe Acute Respiratory Syndrome Coronavirus 2
SHAP: SHapley Additive exPlanations
STAI: State-Trait Anxiety Inventory
STAIS: State anxiety
STAIT: Trait anxiety
XAI: eXplainable Artificial Intelligence

## Acknowledgements

This work was supported by São Paulo Research Foundation (FAPESP grant #2022/11795-3) and by the European Union Horizon 2020 Research and Innovation Programme under ZIKAlliance (grant #734548). MLGK received scholarships from Coordenação de Aperfeiçoamento de Pessoal de Nível Superior (CAPES) - Finance Code 001 and FAPESP (grant #2023/16277-3). CC was supported by FAPESP (grant #2021/00607-9). HB was supported by Conselho Nacional de Desenvolvimento Científico e Tecnológico (CNPq) #310823/2021-8. The Santos Dumont (SDumont) supercomputer was partially involved in the experiments.

FAPESP, CAPES and CNPq had no role in the design and conduct of the study.

## References

Ahmed, R., Fahad, N., Miah, M. S. U., Hossen, Md. J., Morol, Md. K., Mahmud, M., & Mostafizur Rahman, M. (2024). A novel integrated logistic regression model enhanced with recursive feature elimination and explainable artificial intelligence for dementia prediction. Healthcare Analytics, 6, 100362. 10.1016/j.health.2024.100362

Alacevich, C., Thalmann, I., Nicodemo, C., de Lusignan, S., & Petrou, S. (2023). Depression and anxiety during and after episodes of COVID-19 in the community. Scientific Reports, 13(1), 8257. 10.1038/s41598-023-33642-w

Alaedini, A., Lightman, S., & Wormser, G. P. (2024). Is Low Cortisol a Marker of Long COVID? The American Journal of Medicine, 137(7), 564–565. 10.1016/j.amjmed.2024.03.013

Allotey, J., Stallings, E., Bonet, M., Yap, M., Chatterjee, S., Kew, T., Debenham, L., Llavall, A. C., Dixit, A., Zhou, D., Balaji, R., Lee, S. I., Qiu, X., Yuan, M., Coomar, D., Sheikh, J., Lawson, H., Ansari, K., Wely, M. van, … Thangaratinam, S. (2020). Clinical manifestations, risk factors, and maternal and perinatal outcomes of coronavirus disease 2019 in pregnancy: Living systematic review and meta- analysis. BMJ, 370, m3320. 10.1136/bmj.m3320

Ayed, M., Embaireeg, A., Kartam, M., More, K., Alqallaf, M., AlNafisi, A., Alsaffar, Z., Bahzad, Z., Buhamad, Y., Alsayegh, H., Al-Fouzan, W., & Alkandari, H. (2022). Neurodevelopmental outcomes of infants born to mothers with SARS-CoV-2 infection during pregnancy: A national prospective study in Kuwait. 10.21203/rs.3.rs-1209503/v1

Biaggio, A. M. B., Natalício, L., & Spielberger, C. D. (1977). Desenvolvimento da forma experimental em português do Inventário de Ansiedade Traço-Estado (IDATE) de Spielberger. Arquivos brasileiros de psicologia aplicada, 29(3), 31–44.

Bujang, M. A., Sa’at, N., Sidik, T. M. I. T. A. B., & Joo, L. C. (2018). Sample Size Guidelines for Logistic Regression from Observational Studies with Large Population: Emphasis on the Accuracy Between Statistics and Parameters Based on Real Life Clinical Data. The Malaysian Journal of Medical Sciences : MJMS, 25(4), 122–130. 10.21315/mjms2018.25.4.12

Cavazos-Rehg, P. A., Krauss, M. J., Spitznagel, E. L., Bommarito, K., Madden, T., Olsen, M. A., Subramaniam, H., Peipert, J. F., & Jean Bierut, L. (2015). Maternal age and risk of labor and delivery complications. Maternal and child health journal, 19(6), 1202–1211. 10.1007/s10995-014-1624-7

Chen, T., & Guestrin, C. (2016). XGBoost: A Scalable Tree Boosting System. Proceedings of the 22nd ACM SIGKDD International Conference on Knowledge Discovery and Data Mining, 785–794. 10.1145/2939672.2939785

Ciapponi, A., Bardach, A., Comandé, D., Berrueta, M., Argento, F. J., Cairoli, F. R., Zamora, N., María, V. S., Xiong, X., Zaraa, S., Mazzoni, A., & Buekens, P. (2021). COVID-19 and pregnancy: An umbrella review of clinical presentation, vertical transmission, and maternal and perinatal outcomes. PLOS ONE, 16(6), Artigo 6. 10.1371/journal.pone.0253974

Costa, H. da S., Cardoso, A. C., Netto, C. M., Martins-Jr, D. C., & Simões, S. N. (2022). A Framework for prediction of dropout in distance learning through XAI techniques in Virtual Learning Environment. Encontro Nacional de Inteligência Artificial e Computacional (ENIAC), 270–281. 10.5753/eniac.2022.227586

Davis, H. E., McCorkell, L., Vogel, J. M., & Topol, E. J. (2023). Long COVID: Major findings, mechanisms and recommendations. Nature Reviews Microbiology, 21(3), 133–146. 10.1038/s41579-022-00846-2

Engjom, H. M., Ramakrishnan, R., Vousden, N., Bunch, K., Morris, E., Simpson, N., Gale, C., O’Brien, P., Quigley, M., Brocklehurst, P., Kurinczuk, J. J., & Knight, M. (2024). Perinatal outcomes after admission with COVID-19 in pregnancy: A UK national cohort study. Nature Communications, 15(1), 3234. 10.1038/s41467-024-47181-z

Hosseini, S. M., Biglan, M. W., Larkby, C., Brooks, M. M., Gorin, M. B., & Day, N. L. (2009). Trait anxiety in pregnant women predicts offspring birth outcomes. Paediatric and Perinatal Epidemiology, 23(6), 557–566. 10.1111/j.1365-3016.2009.01065.x

Huang, J.-C., Lyu, S.-C., Pan, B., Wang, H.-X., Ma, Y.-W., Jiang, T., He, Q., & Lang, R. (2025). A logistic regression model to predict long-term survival for borderline resectable pancreatic cancer patients with upfront surgery. Cancer Imaging, 25(1), 10. 10.1186/s40644-025-00830-y

Johnson, R. C., & Slade, P. (2003). Obstetric complications and anxiety during pregnancy: Is there a relationship? Journal of Psychosomatic Obstetrics & Gynecology. 10.3109/01674820309042796

Ke, G., Meng, Q., Finley, T., Wang, T., Chen, W., Ma, W., Ye, Q., & Liu, T.-Y. (2017). LightGBM: A highly efficient gradient boosting decision tree. Proceedings of the 31st International Conference on Neural Information Processing Systems, 3149–3157.

Khalil, A., Syngelaki, A., Maiz, N., Zinevich, Y., & Nicolaides, K. H. (2013). Maternal age and adverse pregnancy outcome: A cohort study. Ultrasound in Obstetrics & Gynecology: The Official Journal of the International Society of Ultrasound in Obstetrics and Gynecology, 42(6), 634–643. 10.1002/uog.12494

Klein, J., Wood, J., Jaycox, J. R., Dhodapkar, R. M., Lu, P., Gehlhausen, J. R., Tabachnikova, A., Greene, K., Tabacof, L., Malik, A. A., Silva Monteiro, V., Silva, J., Kamath, K., Zhang, M., Dhal, A., Ott, I. M., Valle, G., Peña-Hernández, M., Mao, T., … Iwasaki, A. (2023). Distinguishing features of long COVID identified through immune profiling. Nature, 623(7985), 139–148. 10.1038/s41586-023-06651-y

Knobel, R., Lopes, T. J. P., Menezes, M. de O., Andreucci, C. B., Gieburowski, J. T., & Takemoto, M. L. S. (2020). Cesarean-section Rates in Brazil from 2014 to 2016: Cross-sectional Analysis Using the Robson Classification. Revista Brasileira de Ginecologia e Obstetrícia, 42, 522–528. 10.1055/s-0040-1712134

Luft, C. D. B., Sanches, S. de O., Mazo, G. Z., & Andrade, A. (2007). Versão brasileira da Escala de Estresse Percebido: Tradução e validação para idosos. Revista de Saúde Pública, 41, 606–615. 10.1590/S0034-89102007000400015

Lundberg, S. M., & Lee, S.-I. (2017). A Unified Approach to Interpreting Model Predictions. Advances in Neural Information Processing Systems, 30. https://proceedings.neurips.cc/paper/2017/hash/8a20a8621978632d76c43dfd28b67 767-Abstract.html

Monod, M., Blenkinsop, A., Xi, X., Hebert, D., Bershan, S., Tietze, S., Baguelin, M., Bradley, V. C., Chen, Y., Coupland, H., Filippi, S., Ish-Horowicz, J., McManus, M., Mellan, T., Gandy, A., Hutchinson, M., Unwin, H. J. T., Elsland, S. L. van, Vollmer, M. A. C., … Team, on behalf of the I. C. C.-19 R. (2021). Age groups that sustain resurging COVID-19 epidemics in the United States. Science (New York, N.y.), eabe8372. 10.1126/science.abe8372

Morrow, E. L., Duff, M. C., & Mayberry, L. S. (2022). Mediators, Moderators, and Covariates: Matching Analysis Approach for Improved Precision in Cognitive-Communication Rehabilitation Research. Journal of Speech, Language, and Hearing Research : JSLHR, 65(11), 4159. 10.1044/2022_JSLHR-21-00551

Okeke, S. R., Idriss-Wheeler, D., & Yaya, S. (2022). Adolescent pregnancy in the time of COVID-19: What are the implications for sexual and reproductive health and rights globally? Reproductive Health, 19(1), 207. 10.1186/s12978-022-01505-8

Pedregosa, F., Varoquaux, G., Gramfort, A., Michel, V., Thirion, B., Grisel, O., Blondel, M., Prettenhofer, P., Weiss, R., Dubourg, V., Vanderplas, J., Passos, A., Cournapeau, D., Brucher, M., Perrot, M., & Duchesnay, É. (2011). Scikit-learn: Machine Learning in Python. The Journal of Machine Learning Research, 12(85), 2825–2830. 10.5555/1953048.2078195

Peduzzi, P., Concato, J., Kemper, E., Holford, T. R., & Feinstein, A. R. (1996). A simulation study of the number of events per variable in logistic regression analysis. Journal of Clinical Epidemiology, 49(12), 1373–1379. 10.1016/S0895-4356(96)00236-3

Prokhorenkova, L., Gusev, G., Vorobev, A., Dorogush, A. V., & Gulin, A. (2019). CatBoost: Unbiased boosting with categorical features (arXiv:1706.09516). arXiv. 10.48550/arXiv.1706.09516

Rejnö, G., Lundholm, C., Öberg, S., Lichtenstein, P., Larsson, H., D’Onofrio, B., Larsson, K., Saltvedt, S., Brew, B. K., & Almqvist, C. (2019). Maternal anxiety, depression and asthma and adverse pregnancy outcomes – a population based study. Scientific Reports, 9(1), 13101. 10.1038/s41598-019-49508-z

Rosseel, Y. (2012). lavaan: An R Package for Structural Equation Modeling. Journal of Statistical Software, 48, 1–36. 10.18637/jss.v048.i02

São Paulo Public Security Secretariat. (2024). SP Dados Criminais (Versão 2024) [XLSX]. SP DADOS - RES 160 - RES 516; https://www.ssp.sp.gov.br/assets/estatistica/transparencia/spDados/SPDadosCrimin ais_2024.xlsx. https://www.ssp.sp.gov.br/estatistica/consultas

Taquet, M., Sillett, R., Zhu, L., Mendel, J., Camplisson, I., Dercon, Q., & Harrison, P. J. (2022). Neurological and psychiatric risk trajectories after SARS-CoV-2 infection: An analysis of 2-year retrospective cohort studies including 1 284 437 patients. The Lancet Psychiatry, 9(10), 815–827. 10.1016/S2215-0366(22)00260-7

World Health Organization. (2024). Weekly epidemiological update on COVID-19—17 *May* 2024 (Epidemiological Report 167; Emergency Situation Updates, p. 28). World Health Organization. https://www.who.int/publications/m/item/weekly-epidemiological-update-on-covid-19 16-march-2023

Yang, H., Chen, W., Hu, Y., Chen, Y., Zeng, Y., Sun, Y., Ying, Z., He, J., Qu, Y., Lu, D., Fang, F., Valdimarsdóttir, U. A., & Song, H. (2020). Pre-pandemic psychiatric disorders and risk of COVID-19: A UK Biobank cohort analysis. The Lancet Healthy Longevity, 1(2), e69–e79. 10.1016/S2666-7568(20)30013-1

Zulaika, G., Bulbarelli, M., Nyothach, E., van Eijk, A., Mason, L., Fwaya, E., Obor, D., Kwaro, D., Wang, D., Mehta, S. D., & Phillips-Howard, P. A. (2022). Impact of COVID-19 lockdowns on adolescent pregnancy and school dropout among secondary schoolgirls in Kenya. BMJ Global Health, 7(1), e007666. 10.1136/bmjgh-2021-007666

