## Supplementary Table 1 for "COVID-19 modulates pregnancy outcomes"

**Table S1 - Potential exposure variables used in the study.**

| Variable name | Variable type | Values | Methodology and sources | % missing |
|---|---|---|---|---|
| Stress in the last 30 days | Quantitative | Natural numbers.<br>Ranges from 0 to 40. | Measured by the 10-item Perceived Stress Scale (PPS-10) [1], translated to Brazilian Portuguese and validated in a Brazilian sample [2]. | 6.50% |
| Stressful events during lifetime | Quantitative | Natural numbers.<br>Ranges from 0 to 12. | Measured by an in-house questionnaire with 12 yes/no questions. Examples of questions are death of a relative, residence eviction, and contact with child protective services. | 4.07% |
| Stressful events during gestation | Quantitative | Natural numbers.<br>Ranges from 0 to 6. | Measured by an in-house with 6 yes/no questions. Examples of questions are fight with a friend, residence eviction, and violence. | 4.88% |
| State anxiety | Quantitative | Natural numbers.<br>Ranges from 0 to 80. | Measured by the 20-item State Anxiety Inventory [3,4], form X, translated to Brazilian Portuguese and validated in a Brazilian sample [4]. | 5.69% |
| Trait anxiety | Quantitative | Natural numbers.<br>Ranges from 0 to 80. | Measured by the 20-item Trait Anxiety Inventory [3], form X, translated to Brazilian Portuguese and validated in a Brazilian sample [4]. | 3.25% |
| Weight gain during pregnancy | Qualitative - Ordinal | 0 - Less than 8kg<br>1 - Between 8kg and 12kg<br>2 - Between 12kg and 18kg<br>3 - More than 18kg | Interview and evaluation at delivery at HU-USP. | 2.44% |
| Prenatal BMI | Quantitative | Any positive rational number, usually ranges between 15 and 45. | Interview at delivery at HU-USP. | 4.07% |
| Prenatal appointments | Quantitative | Natural numbers.<br>Ranges from 0 to 9. More than 9 appointments were not accounted. | Interview and evaluation at delivery at HU-USP. | 0.81% |

| | | | | |
|---|---|---|---|---|
| Intentional violent crime rate | Quantitative | Any positive rational number. Number of intentional violent crimes per ten thousand inhabitants. | In the interview at delivery, we collected the patient's zip code. Using the Brazilian Post and Telegraph Corporation database, we mapped them to their municipality. If the municipality was São Paulo, we further mapped them to one of the 96 city districts.<br><br>We calculated the violent crime rate with our Python module SPCrime.The base year was 2022, which corresponds to the earliest data publicly provided by the São Paulo Secretariat of Public Security [5], and closest to our sample recruitment timeframe. The population size data came from the Brazilian Institute of Geography and Statistics (IBGE) census of 2022 [6].<br><br>Brazilian security organs consider the following categories as intentional violent crimes [7]: intentional homicide, armed robbery resulting in death ('latrocínio'), bodily injury followed by death, rape (both of vulnerable person or not), intentional bodily injury, attempted murder. | 9.76% |
| Maternal age | Quantitative | Natural numbers.<br>In our sample, the range is 15 to 41 | Interview at delivery at HU-USP. | 0% |
| Paternal age | Quantitative | Natural numbers.<br>In our sample, the range is 18 to 69 | Interview at delivery at HU-USP. | 4.88% |
| Income | Quantitative | Natural numbers in BRL (Real)<br>In our sample, the range is 0 to 8000 | Interview at delivery at HU-USP. | 14.63% |
| Maternal education | Qualitative - Ordinal | 0 - Illiterate<br>1 - Incomplete elementary/middle school (1st to 9th grade)<br>2 - Complete elementary/middle school (1st to 9th grade) | Interview at delivery at HU-USP.<br><br>"Incomplete" may mean either dropout or ongoing. | 0% |

| | | | | |
|---|---|---|---|---|
| | | 3 - Incomplete high school (10th to 12th grade)<br>4 - Complete high school (10th to 12th grade)<br>5 - Incomplete college education<br>6 - Complete college education<br>7 - Post-graduate studies<br><br>There were no 0 or 7 values in our sample. | | |
| Racial or ethnic minority | Qualitative - Boolean | 0 - White or East Asian people<br>1 - Black, Mixed or Indigenous people | Interview at delivery at HU-USP.<br><br>We considered the five categories of the IBGE [6]. Here we translated them: 'Branco' as 'White people', 'Amarelo' as 'East Asian people', 'Preto' as 'Black people', 'Pardo' as 'Mixed-race people', and 'Indígena' as 'Indigenous people'.<br><br>Although other countries may consider East Asian people as a racial minority, in Brazil they are not, and do not face the same challenges as Black, Mixed and Indigenous people. | 0% |
| Worker or student | Qualitative - Boolean | 0 - Unemployed or homeowner. | Interview at delivery at HU-USP. | 0% |

**References**


[1] S. Cohen, T. Kamarck, R. Mermelstein, A Global Measure of Perceived Stress, Journal of Health and Social Behavior 24 (1983) 385–396. https://doi.org/10.2307/2136404.

[2] C.D.B. Luft, S. de O. Sanches, G.Z. Mazo, A. Andrade, Versão brasileira da Escala de Estresse Percebido: tradução e validação para idosos, Rev. Saúde Pública 41 (2007) 606–615. https://doi.org/10.1590/S0034-89102007000400015.

[3] C.D. Spielberger, F. Gonzalez-Reigosa, A. Martinez-Urrutia, L.F. Natalicio, D.S. Natalicio, The state-trait anxiety inventory, Revista Interamericana de Psicologia/Interamerican Journal of Psychology 5 (1971).

[4] A.M.B. Biaggio, L. Natalício, C.D. Spielberger, Desenvolvimento da forma experimental em português do Inventário de Ansiedade Traço-Estado (IDATE) de Spielberger, Arquivos Brasileiros de Psicologia Aplicada 29 (1977) 31–44.

[5] São Paulo Public Security Secretariat, SP Dados Criminais, (2024). https://www.ssp.sp.gov.br/estatistica/consultas (accessed June 18, 2024).

[6] Brazilian Institute for Geography and Statistics, Censo Demográfico 2022 : população e domicílios : primeiros resultados / IBGE, Coordenação Técnica do Censo Demográfico, 1st ed., Brazilian Institute for Geography and Statistics, Rio de Janeiro, 2023. https://biblioteca.ibge.gov.br/index.php/biblioteca-catalogo?view=detalhes&id=2102011 (accessed September 13, 2024).

[7] E.H.B. Oliveira, Metodologia para aferição de indicadores e metasda secretaria de segurança pública de Goiás, Secretaria de Segurança Pública do Estado de Goiás, Goiânia, 2019. https://www.seguranca.go.gov.br/wp-content/uploads/2019/04/portaria-n-0236-19-ssp-de-auditoria-abril-2019-2.pdf (accessed June 15, 2024).
