## Supplementary Table 2 for "COVID-19 modulates pregnancy outcomes"

**Table S2.** Value grid explored in the hyperparameter tuning of the classifier models. The search, performed with the function RandomizedSearchCV() from Scikit-Learn, explored 100 random combinations at each loop.

| Model | N° combinations | Hyperparameter | Values |
|---|---|---|---|
| **RandomForestClassifier** | **6912** | max_depth | None, 2, 3, 4, 5, 10, 20, 30 |
| | | min_samples_leaf | 2, 4, 5, 7 |
| | | min_samples_split | 2, 5, 10 |
| | | n_estimators | 50, 100, 200, 600, 1000, 1500 |
| | | criterion | gini, entropy, log_loss |
| | | bootstrap | True, False |
| | | max_features | sqrt, log2 |
| **LogisticRegression** | **60** | solver | lbfgs, newton-cholesky, liblinear |
| | | class_weight | None, balanced |
| | | penalty | ['l2', None] if solver lbfgs or newton-cholesky, ['l1', 'l2'] if solver = liblinear |
| | | C | 0.1, 1, 10, 100, 1000 |
| **LGBMClassifier** | **57600** | boosting_type | dart, gbdt |
| | | max_bin | 50, 150, 255, 300 |
| | | max_depth | 3, 4, 5, 6 |
| | | num_leaves | 2^max_depth |
| | | min_data_in_leaf | 3, 5, 7, 10, |
| | | lambda_l1 | 1, 0.1, 0.01, 0.0001, 0 |
| | | lambda_l2 | 1, 0.1, 0.01, 0.0001, 0 |

| | | | |
|---|---|---|---|
| | | feature_fraction | 0.5, 0.8, 1 |
| | | num_iterations | 50, 100, 150 |
| | | learning_rate | [0.005, 0.01] if num_iterations = 150, [0.025, 0.05] if num_iterations = 100,<br>[0.1, 0.2] if num_iterations = 50 |
| | | extra_trees | True |
| **XGBClassifier** | **54000** | alpha | 1, 0.1, 0.01, 0.0001, 0 |
| | | colsample_bytree | 0.6, 0.8, 1.0 |
| | | eta | 0.01, 0.05, 0.1, 0.2 |
| | | gamma | 0, 0.5, 1, 2, 5 |
| | | lambda | 1, 0.1, 0.01, 0.0001, 0 |
| | | max_depth | 3, 4, 5, 6 |
| | | min_child_weight | 3, 6, 10 |
| | | subsample | 0.6, 0.8, 1.0 |
| **CatBoostClassifier** | **30000** | iterations | 50, 100, 150, 300, 500 |
| | | depth | 3, 4, 5, 6, 7, 8, 9, 10 |
| | | loss_function | Logloss, CrossEntropy |
| | | l2_leaf_reg | 1, 0.1, 0.01, 0.0001, 0 |
| | | leaf_estimation_iterations | 1, 3, 5 |
| | | learning_rate | 0.005, 0.01, 0.05, 0.1, 0.2 |
| | | min_child_samples | 1, 4, 8, 16, 32 |
| **SVC** | **75** | C | 0.1, 1, 10, 100, 1000 |

| | | | |
|---|---|---|---|
| | | gamma | 1, 0.1, 0.01, 0.001, 0.0001, |
| | | kernel | rbf, linear, poly |
| **KNeighborsClassifier** | **174** | n_neighbors | 2, 3, 4, 5, 6, 7, 8, 9, 10, 11, 12, 13, 14, 15, 16, 17, 18, 19, 20, 21, 22, 23, 24, 25, 26, 27, 28, 29, 30 |
| | | weights | uniform, distance |
| | | algorithm | ball_tree, kd_tree, brute |
| **GaussianNB** | **100** | var_smoothing | numpy.logspace(0,-9, num=100) |
