## Supplementary Table 3 for "COVID-19 modulates pregnancy outcomes"

**Supplementary Table 3 -** Sample description. The table presents key demographic and mental health characteristics. The full dataset is available as Supplementary Material 1. Quantitative variables are represented as average ± standard deviation. Qualitative variables are summarized as percentages.

| | | **CONTROL (n = 77)** | **COVID (n = 43)** |
|---|---|---|---|
| | **Maternal age (years)** | 28.71 ± 6.73 | 26.49 ± 6.16 |
| **Maternal educational level** | Elementary School (ongoing or dropout) | 6 (7.79%) | 0( 0.00%) |
| | Elementary School | 10 (13.00%) | 3 (6.98%) |
| | High School (ongoing or dropout) | 10 (12.99%) | 10 (23.26%) |
| | High School | 34 (44.16%) | 25 (58.14%) |
| | College (ongoing or dropout) | 7 (9.09%) | 1 (2.33%) |
| | College | 10 (12.99%) | 4 (9.30%) |
| | **Income (BRL)** | 2623.16 ±1487.23 | 2314.29±1106.70 |
| | **Prenatal BMI** | 26.13±5.47 | 27.62±5.88 |
| | **Racial and ethnic minority (% yes)** | 53 (68.82%) | 25 (58.13%) |
| **Occupation** | Worker or student | 38 (49.35%) | 29 (67.44%) |
| | Homeowner or unemployed | 39 (50.65%) | 14 (32.56%) |
| | **Stress in the last 30 days (PSS10)** | 17.92± 7.10 | 18.02 ± 7.81 |
| | **State anxiety (STAI-S)** | 39.54±9.14 | 39.88±8.42 |
| | **Trait anxiety (STAI-T)** | 37.25±10.76 | 41.38±9.69 |
| **Delivery type** | Cesarean or forceps | 58 (75.33%) | 33 (76.74%) |
| | Vaginal | 19 (24.68%) | 10 (23.26%) |
| | **Preterm (< 37 weeks)** | 10 (14.28%) | 3 (7.14%) |
| | **Neonatal weight at birth (grams)** | 3326.66 ±483.863 | 3297.86±405.96 |

*Legend: BRL - Brazilian real; BMI - Body Mass Index; PSS10 - Perceived Stress Scale (ten-item version).*
