## Supplementary Table 4 for "COVID-19 modulates pregnancy outcomes"

**Supplementary Table 4.** Parameter values and evaluation index for the Moderation Analyses. The columns represent each analysis with a different exposure variable. In the four analyses, the moderator was COVID-19 and the outcome was delivery complications. Asterisks indicate statistical significance.

| | | State anxiety | Trait anxiety | Maternal age | Paternal age |
|---|---|---|---|---|---|
| $\beta_1$ (exposure) | Mean±SD | 0.106±0.04 | 0.146±0.05 | -0.094±0.05 | -0.093±0.04 |
| | p-value | 0.009* | 0.002* | 0.044* | 0.030* |
| $\beta_2$ (COVID-19) | Mean±SD | 0.106±0.09 | 0.051±0.09 | 0.057±0.09 | 0.140±0.09 |
| | p-value | 0.240 | 0.547 | 0.504 | 0.108 |
| $\beta_3$ (exposure·COVID19) | Mean±SD | 0.172±0.07 | 0.231±0.08 | -0.276±0.06 | -0.176±0.05 |
| | p-value | 0.021* | 0.005* | 0.000* | 0.002* |
| $\chi$2 test | df | 2 | 2 | 2 | 2 |
| | p-value | 0.000* | 0.000* | 0.000* | 0.01* |
| $R^2$ | | 0.102 | 0.175 | 0.158 | 0.139 |

*Legend: df = degrees of freedom; SD = standard deviation.*
