## Supplementary Figure 1 for "COVID-19 modulates pregnancy outcomes"

**Supplementary Figure 1 –** Association between potential exposure variables. **a)** Pearson correlation coefficient between the quantitative variables. All correlation coefficients are below 0.7. **b)** p-value after chi-squared test between the qualitative variables. There are no p-values under the significance cut-off of 0.05.

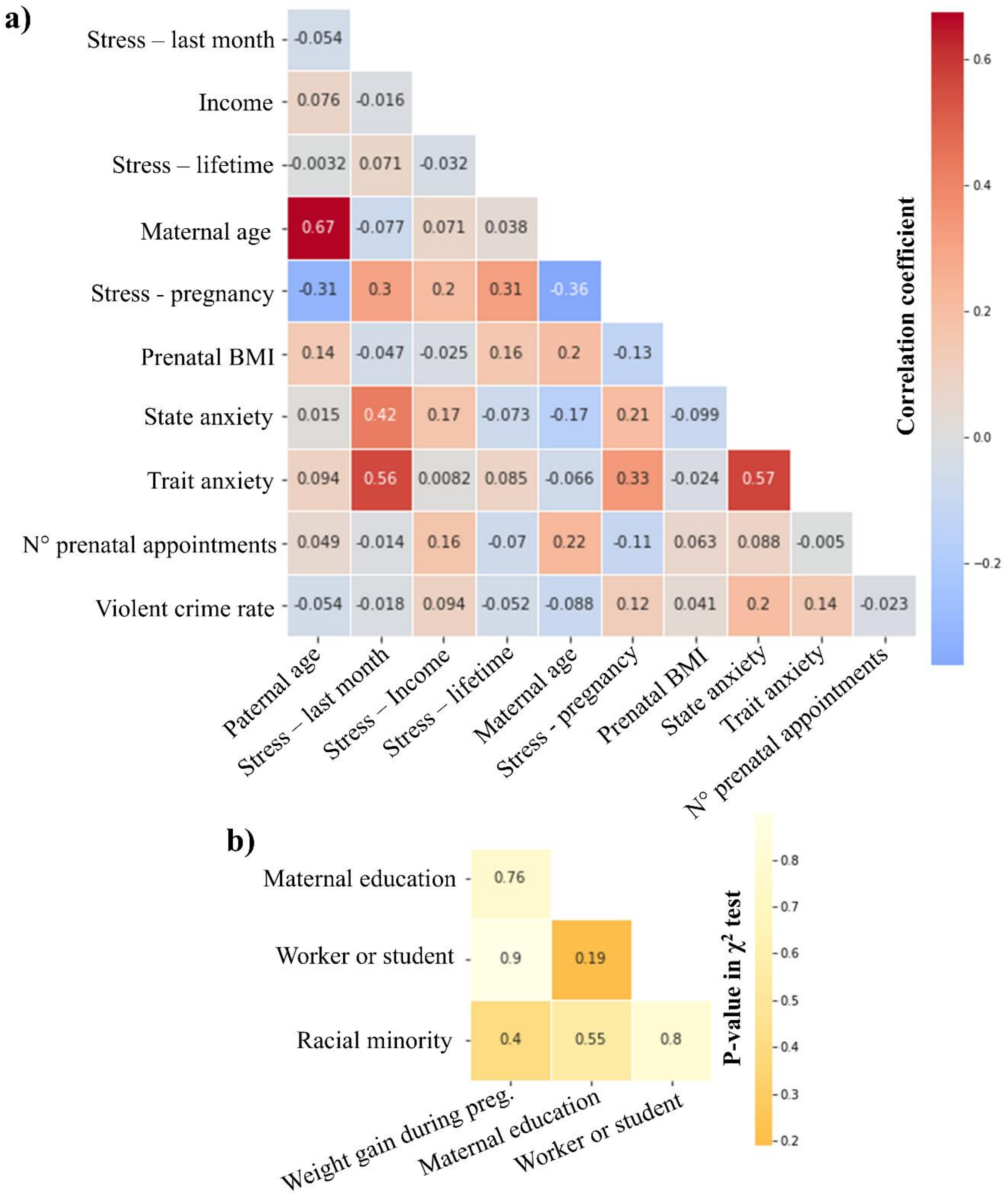
