## Supplementary Figure 2 for "COVID-19 modulates pregnancy outcomes"

**Supplementary figure 2 –** Determination of the best K value in the K-nearest neighbours data imputation. We compared the K values: 3, 4, 5, 7 and 10. **a)** Data distribution of the quantitative variables before and after imputation. Values are standardized by z-score. **b-c)** F1-score and balanced accuracy for a XGBoost model trained with imputed and non-imputed data. XGBoost ran with its default hyperparameters. We used a 80/20 cross-validation scheme repeated 10 times.

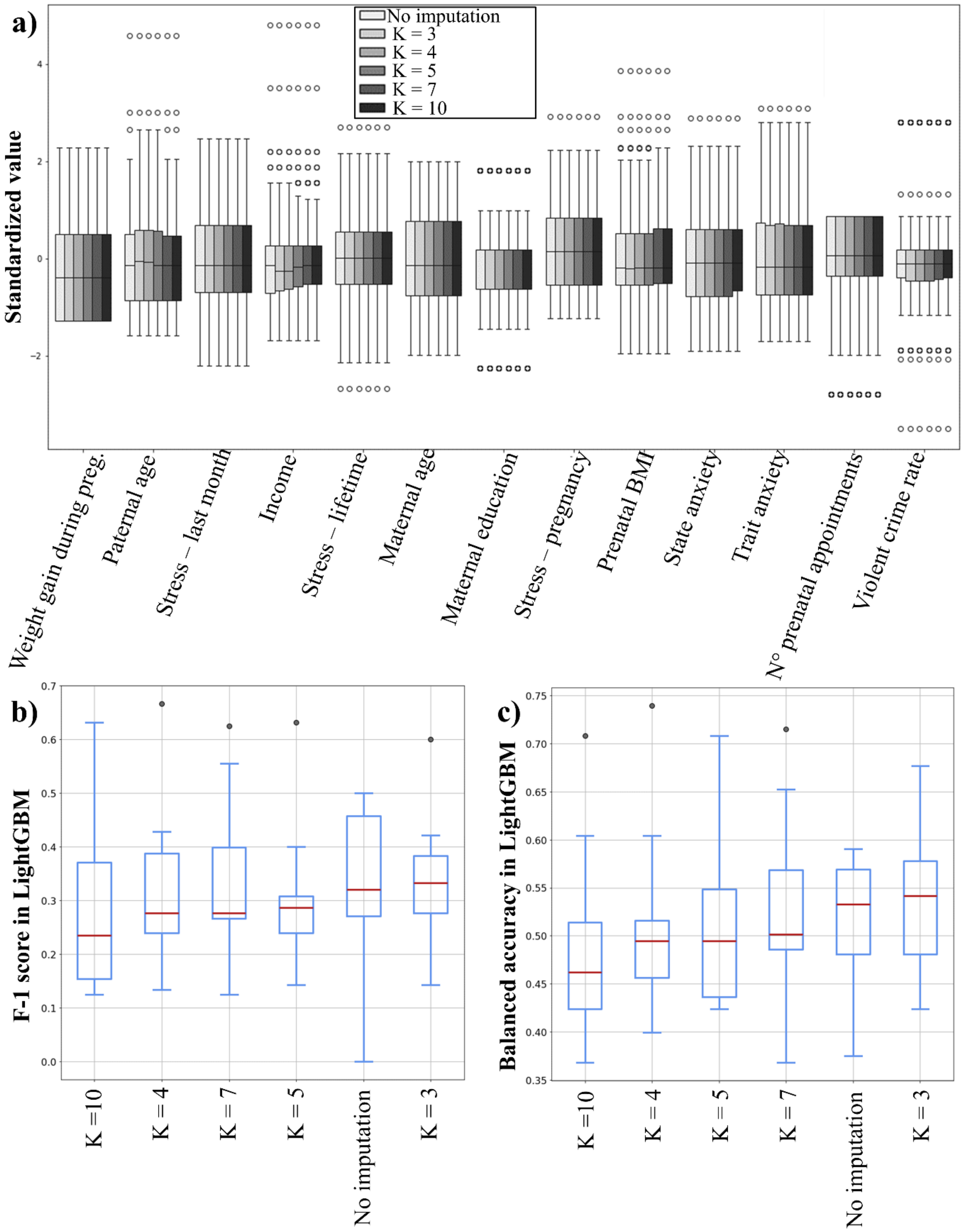
