## Supplementary figures and images for "COVID-19 modulates pregnancy outcomes"

### Supplementary Figure 3

**Supplementary Figure 3** – Flow chart of patient selection.

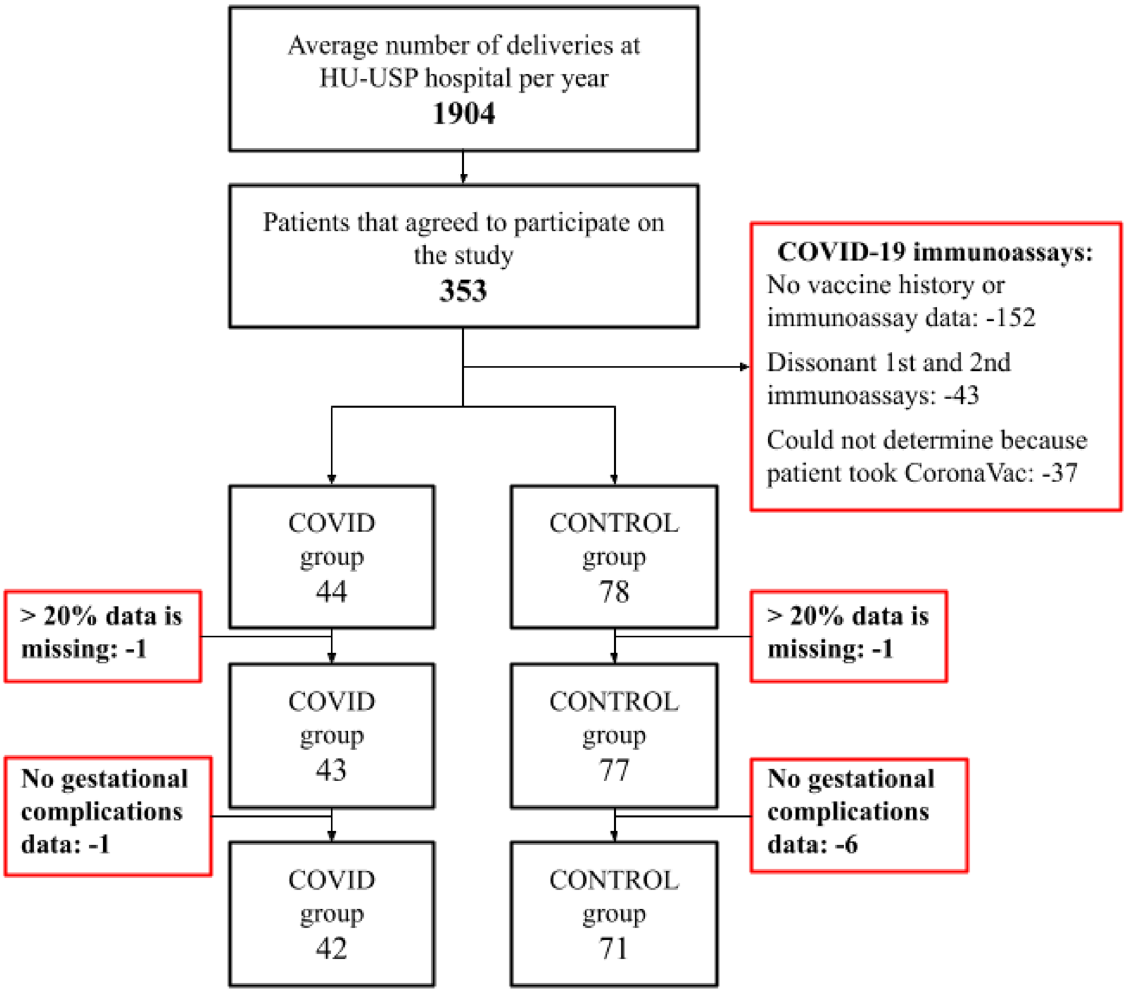
