## Supplementary Figure 4 for "COVID-19 modulates pregnancy outcomes"

**Supplementary Figure 4 –** Pairwise distribution of nine predictor variables. A) Paternal age, B) Stress in the last month, C) Income, D) Maternal Age, E) Prenatal BMI, F) State anxiety, G) Trait Anxiety, H) Number of prenatal appointments, I) violent crime rate at the district of residence of the patient. Blue indicates patients from the CONTROL group while orange indicates patients from the COVID group.

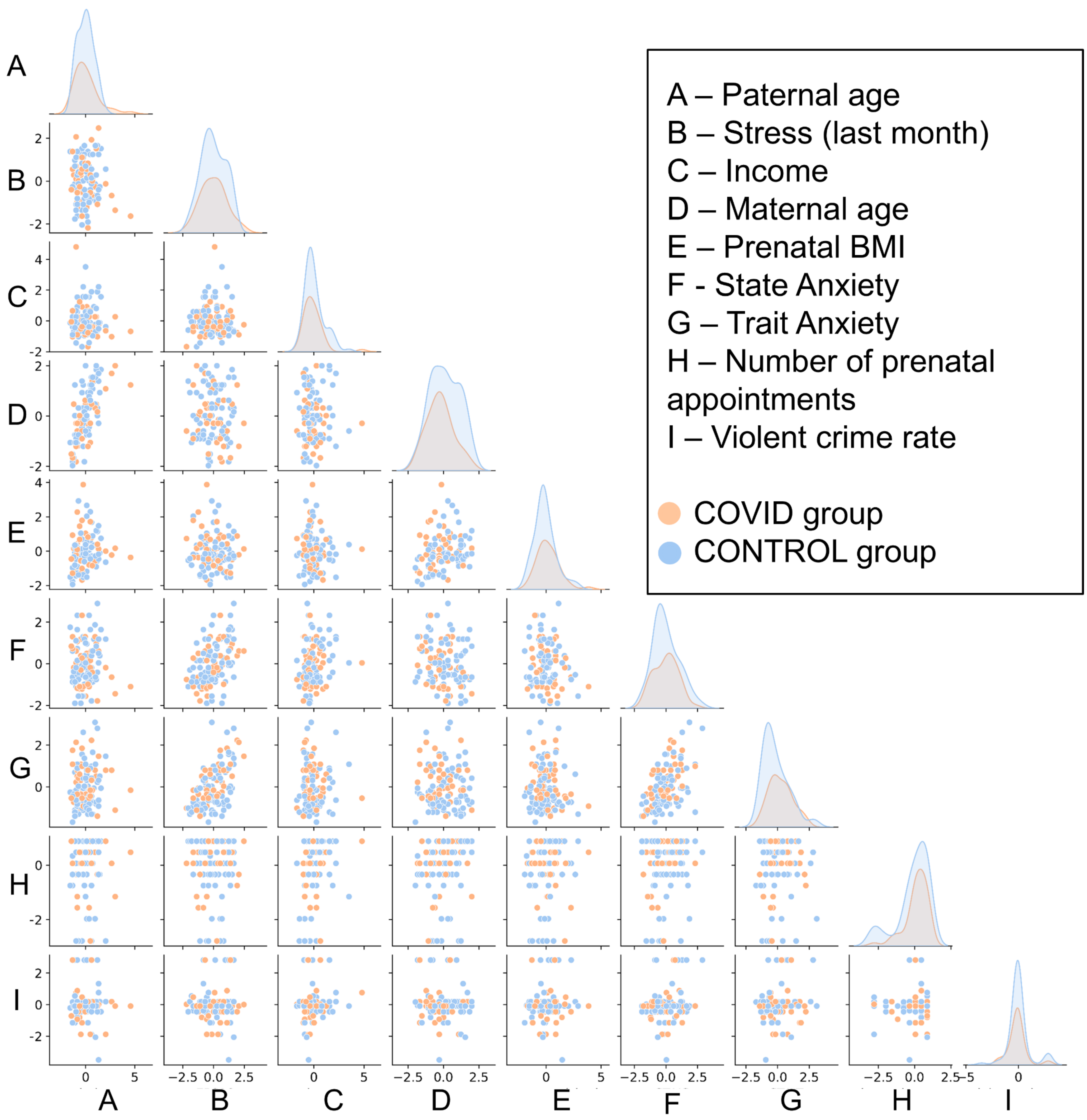
