## Supplementary Figure 5 for "COVID-19 modulates pregnancy outcomes"

**Figure S5 -** Comparison of models predicting whether the pregnant person had COVID-19. Balanced accuracy, F1-score area under the receiver operating characteristic curve (ROC-AUC) and and Jaccard index for the models: Logistic regression and support vector machine (SVM). We fixed three kernels for the SVM algorithm: Radial Basis Function (RBF), polynomial (poly) and linear. Boxes represent the Q2 and Q3 quartiles, the line represents the median, and the whiskers represent 150% of the interquartile range.

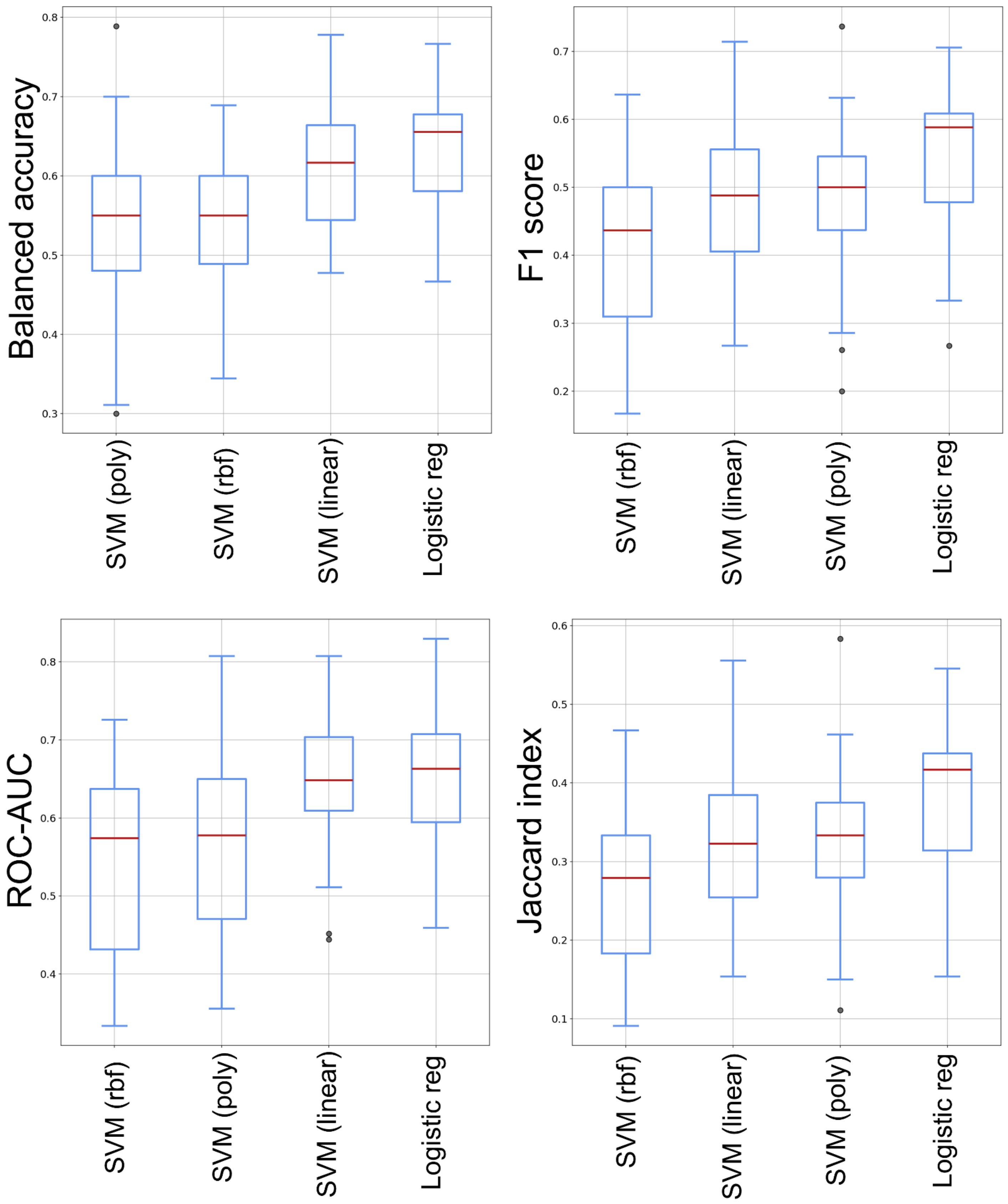
