## Supplementary Figure 6 for "COVID-19 modulates pregnancy outcomes"

**Figure S4 –** Univariate logistic regression where the delivery complications are the outcomes. We computed separately the models for pregnant people who had COVID-19 (COVID) or did not (CONTROL). The predictors are: A) Trait Anxiety, B) State Anxiety, C) Paternal age, D) Maternal age.

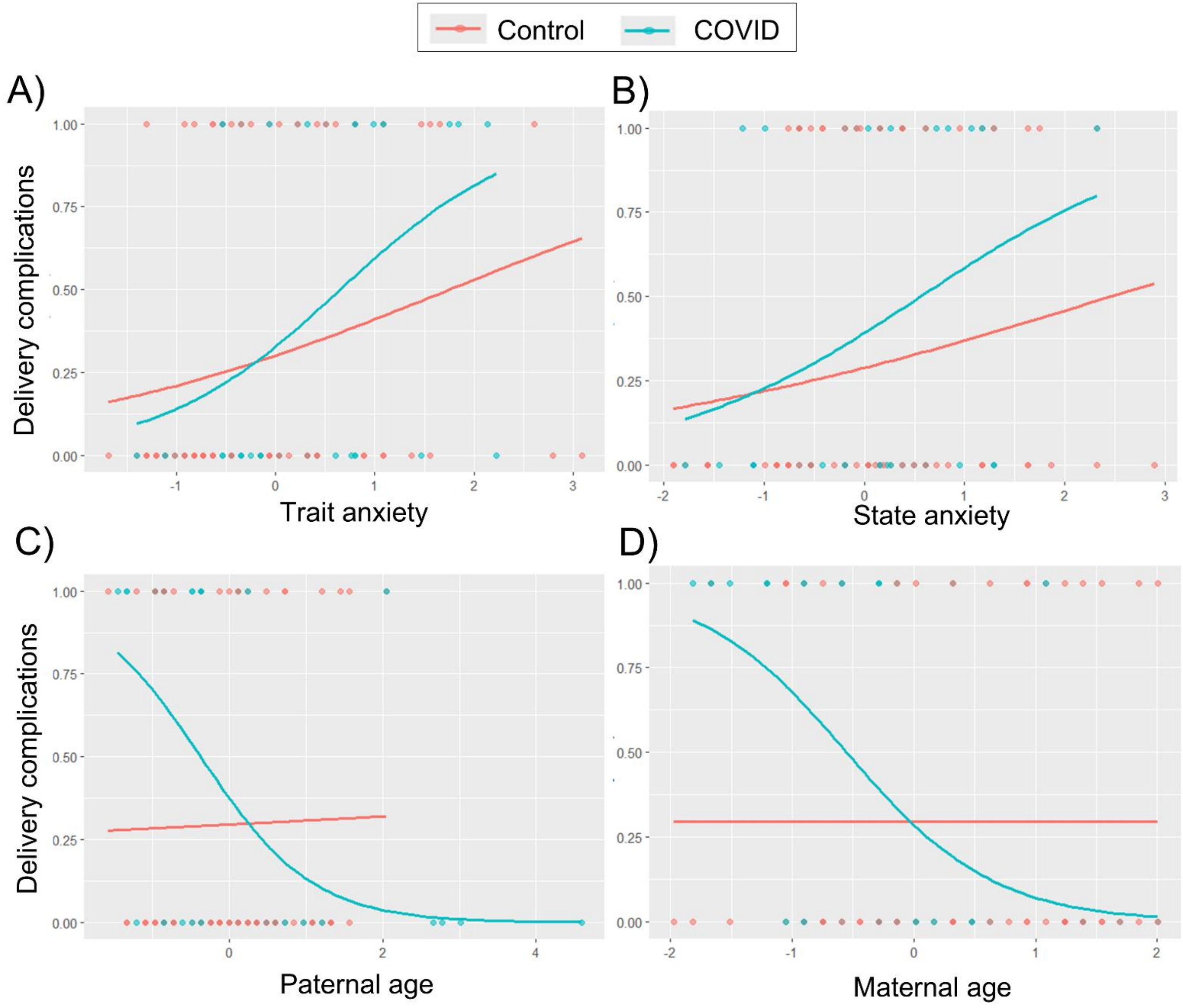
